# Surgical Treatments for Lumbar Spine Diseases: A Systematic Review and Meta-Analysis

**DOI:** 10.1101/2021.07.04.21259978

**Authors:** Kanthika Wasinpongwanich, Tanawin Nopsopon, Krit Pongpirul

**Author notes:** **Corresponding author: Krit Pongpirul**.

## Abstract

**Purpose:** Surgical treatment is mandatory in some patients with lumbar spine diseases. To obtain spine fusion, many operative techniques were developed with different fusion rates and clinical results. This study aimed to collect randomized controlled trial (RCT) data to compare fusion rate, clinical outcomes, complications among Transforaminal Lumbar Interbody Fusion (TLIF), and other techniques for lumbar spine diseases.

**Methods:** A systematic literature search of PubMed, Embase, Scopus, Web of Science, and CENTRAL databases was searched for studies up to 13 February 2020. The meta-analysis was done using a random-effects model. Pooled risk ratio (RR) or mean difference (MD) with a 95% confidence interval of fusion rate, clinical outcomes, and complication in TLIF and other techniques for lumbar diseases.

**Results:** The literature search identified 3,682 potential studies, 15 RCTs (915 patients) were met our inclusion criteria and were included in the meta-analysis. Compared to other techniques, TLIF had slightly lower fusion rate (RR=0.84 [95% CI 0.72, 0.97], *p*=0.02, *I*^*2*^=0.0%) at 1-year follow-up while there was no difference on fusion rate at 2-year follow up (RR=1.06 [95% CI 0.96, 1.18], *p*=0.27, *I*^*2*^=69.0%). The estimated risk ratio of total adverse events (RR=0.90 [95% CI 0.59, 1.38], *p*=0.63, *I*^*2*^=0.0%) and revision rate (RR=0.78 [95%CI 0.34, 1.79], *p*=0.56, *I*^*2*^=39.0%) showed no difference. TLIF had approximately half an hour more operative time than other techniques (MD=31.88 [95% CI 5.33, 58.44], *p*=0.02, *I*^*2*^=92.0%). There was no significant difference between TLIF and other techniques in terms of the blood loss, and clinical outcomes.

**Conclusions:** Besides fusion rate at 1-year follow-up and operative time, our study demonstrated similar outcomes of TLIF with other techniques for lumbar diseases in regard to fusion rate, clinical outcomes, and complications.

## Introduction

Surgical treatment is mandatory in some patients with lumbar spine diseases. To obtain spine fusion, many operative techniques have been developed with different fusion rates and clinical results. Cloward et al. first described Posterolateral Lumbar Interbody Fusion (PLIF) in 1952 [1] whereas Harm & Rollinger introduced Transforaminal Lumbar Interbody Fusion (TLIF) three decades later [2]. In early 2002, the minimally invasive surgical (MIS) approach was promoted to TLIF by Foley and Lefkowiz to improve peri-postoperative morbidity and clinical results [3]. For anterior lumbar interbody fusion has a long history in the tuberculous spine however the technique was adapted to other lumbar spine diseases [4]. Ozgur et al. describe a novel spine procedure called the Extreme Lateral Interbody Fusion or XLIF in 2006 [5].

Several systematic reviews compared either MIS TLIF or Open TLIF with other techniques e.g. MIS vs open TLIF/PLIF [6], TLIF vs ALIF [7], MIS TLIF vs LLIF [8], TLIF vs PLIF [9], TLIF vs PLF [10]. The studies were conducted around 2014-2018 [6-8,10-14]. Most of them compared 1 or 2 techniques with TLIF for lumbar spine diseases [6-14]. Half of them concluded the level of evidence on their study was low and need more randomized control trials (RCTs) [6-8,10,15]. The fusion rates, clinical outcomes, and complications among operative techniques for lumbar spine diseases have been inconclusive. As time passed, the learning curve of different techniques was theoretically decreased and more RCTs were recently propagated[16-30].

This systematic review and meta-analysis aimed to offer updated results based on fusion rate, clinical outcomes, and complications between TLIF, decompression alone (no fusion), posterolateral fusion, and other interbody fusion (PLIF, ALIF, XLIF).

## Methods

This study was conducted following the recommendations of the Preferred Reporting Items of Systematic Reviews and Meta-Analyses (PRISMA) statement. We prospectively registered the systematic review with PROSPERO International Prospective Register of Ongoing Systematic Reviews (Registration number: CRD42020186858).

### Search strategy

A systematic literature search of PubMed, Embase, Scopus, Web of Science, and CENTRAL databases were searched for studies published between January 2010 and January 2019. The electronic databases were searched up to 13 February 2020. The reproducible search strategy was presented in detail in the Supplement. Besides, the reference lists of included articles were searched, as well as related citations from other journals via Google Scholar.

### Study selection

Only randomized controlled trials comparing lumbar interbody fusion with posterolateral fusion and/or other lumbar interbody fusion were anticipated in this review. Inclusion criteria were established as follows: (1) the studies with a population of patient age >18 years old (2) randomized controlled trial investigating lumbar spine disease treated with any Lumbar Interbody Fusion or Posterolateral Fusion or No fusion, (3) the study included at least one outcome (fusion rate, disability and pain or complications, operative time, blood loss, hospital length of stay). Exclusion criteria were (1) biomechanical and cadaveric studies, (2) paper that is not in English, (3) duplicated studies.

The title and abstracts of each study were independently reviewed by 2 authors (KW, TN) to assess for inclusion in the meta-analysis. For studies that meet the inclusion criteria, 2 reviewers (KW, TN) independently reviewed the full manuscripts. Discrepancies between the 2 reviewers were resolved by discussion until reach consensus among the authors. In accordance with PRISMA guidelines, the process is presented in a flow chart [31] (Fig.1)

**Fig. 1.**
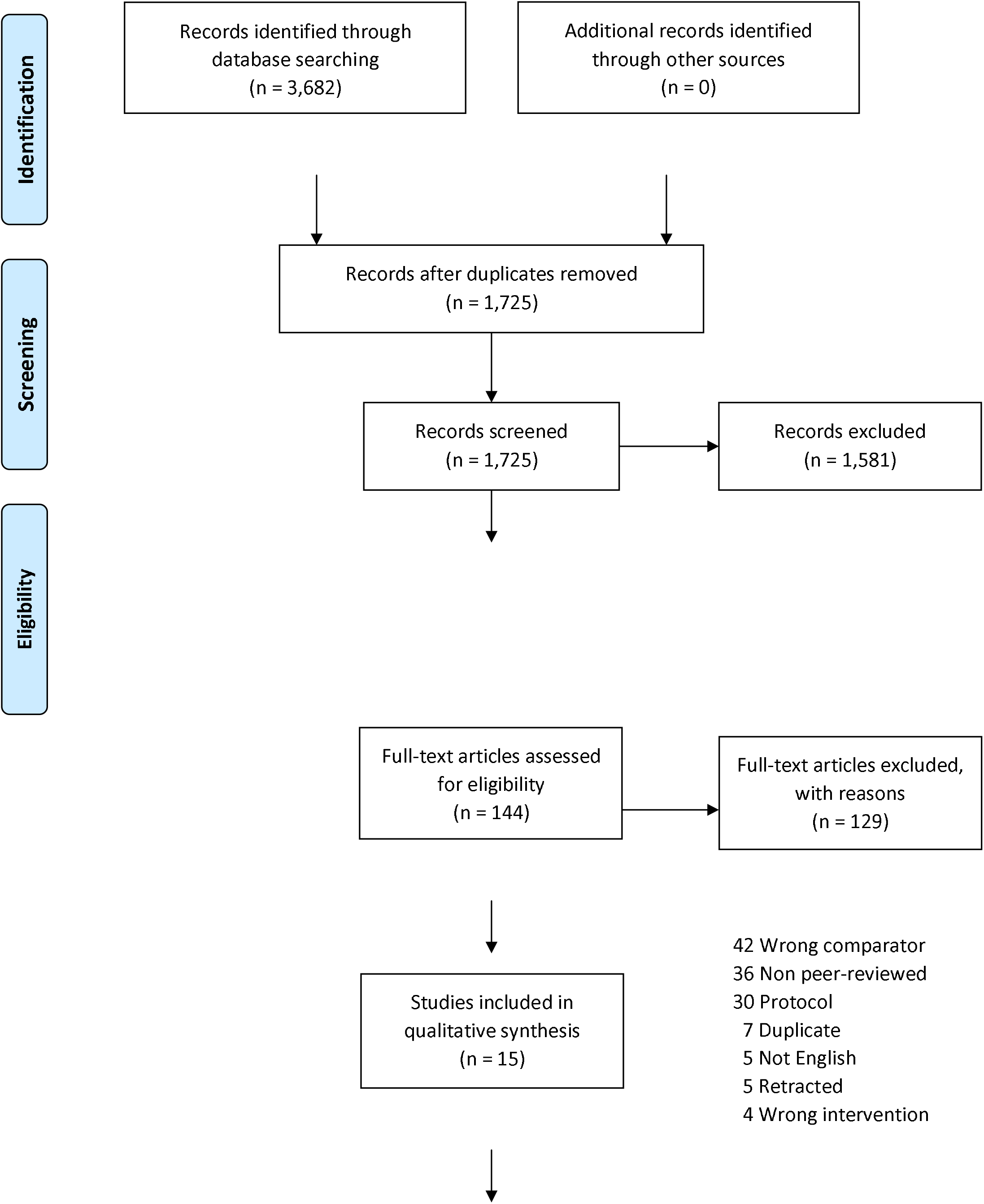
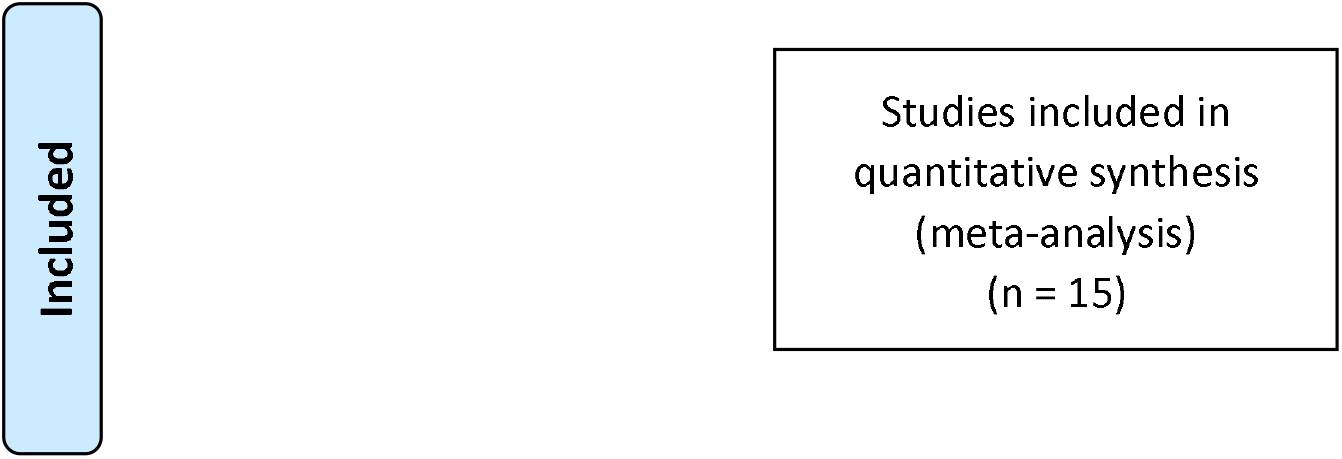
Flowchart of this systematic review with meta-analysis of prospective studies. RCTs, randomized clinical trials

### Data extraction

The following data items were independently extracted by two authors (KW, TN) from the included studies; study design (author, year, country), study population (number of included patients, age, indication for surgery), Visual Analog Score (VAS) for back and leg pain, Oswestry Disability Index (ODI), parameters concerning operation (operative time, length of hospital stay, blood loss, revision) Complications (total adverse events, infection, dural tear, etc.). Discrepancies were resolved by consensus.

### Quality assessment

The authors worked independently to assess the risk of bias in the included trials using the Cochrane Risk of Bias tool 2.0 for a randomized control trial study[32]. We assessed the randomization process, deviations from intended intervention, missing outcome data, measurement of the outcome, selection of the reported result. We assigned each domain as a low risk of bias, some concerns, and a high risk of bias. We contacted the authors if there was not enough information to assess. If the trial authors did not respond within 14 days, we conducted the assessment using available data. We resolved the disagreement through discussion. We presented our risk of bias assessment in Figs. 2 and 3.

**Fig. 2.**
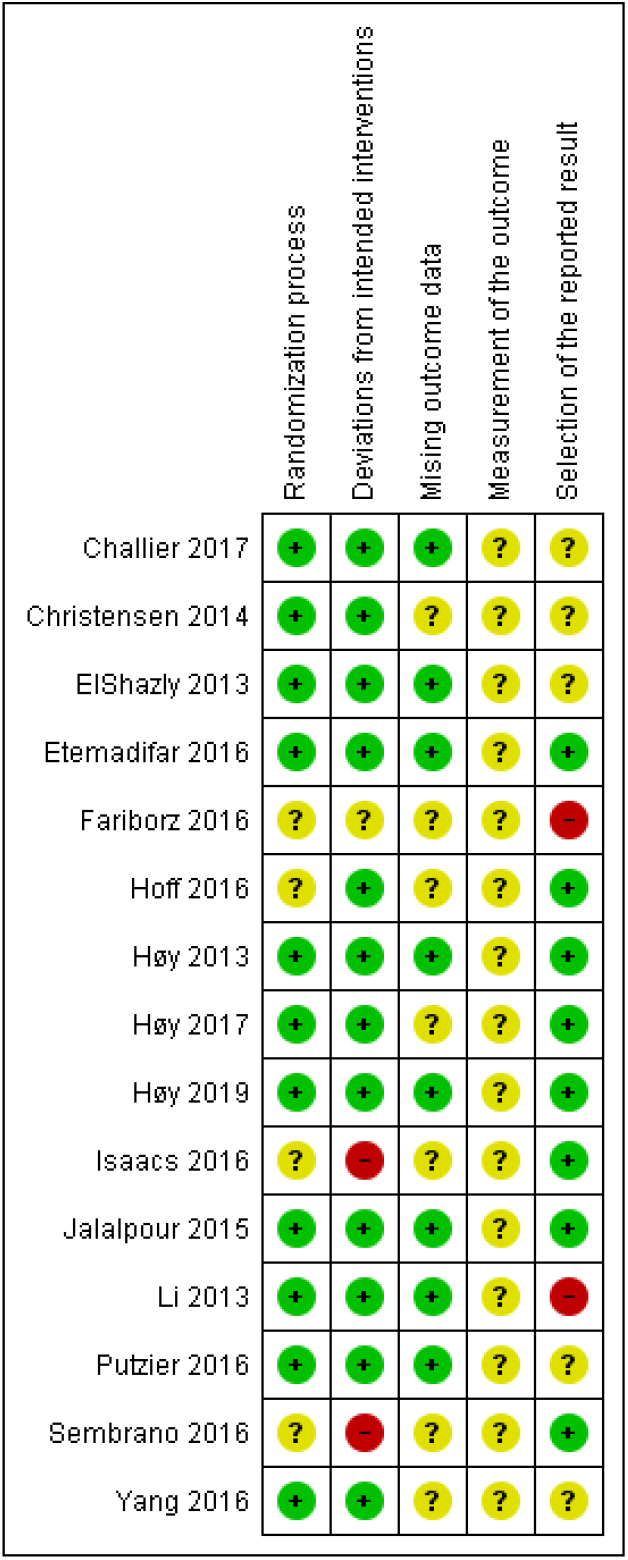
The risk of bias of each included randomized controlled trial. Low risk is presented as green dot, some concerns as yellow dot, and high risk as red dot

**Fig. 3.**
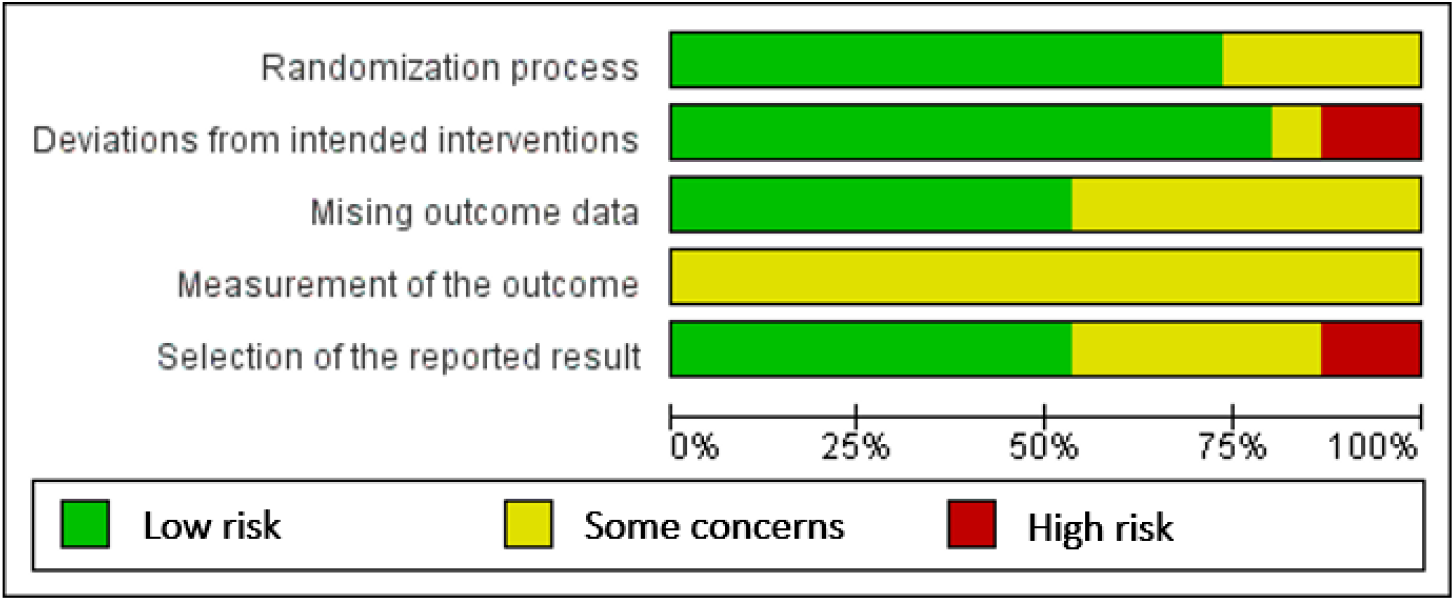
The risk of bias of included randomized controlled trials. Bars show percentages across all included RCTs

### Statistical Analysis

The primary outcome was fusion rate, total adverse events, and revision rate. The outcomes measured were the mean difference for VAS back and leg pain, Oswestry Disability Index (ODI) score, operative time, blood loss, length of hospital stay with associated 95% CI. Fusion rate, total adverse events, infection rate, revision rate, dural tear were reported as the risk ratio (RR) with 95% CI. The results of the studies were included in the meta-analysis and presented in a forest plot, which also showed statistical powers, confidence intervals, and heterogeneity. The variability within-a study and between studies was assessed by an *I*^*2*^ estimate of heterogeneity. We regarded level of heterogeneity for *I*^*2*^ statistic as defined in chapter 9 of the Cochrane Handbook for Systematic Reviews of Interventions: 0–40% might not be important; 30–60% may represent moderate heterogeneity; 50–90% may represent substantial heterogeneity; 75–100% considerable heterogeneity. The random-effects meta-analysis by DerSimonian and Laird method was used as clinical, methodological, and statistical heterogeneity encountered. Prespecified subgroup analyses by type of comparators were performed. We assessed publication bias by computing each study effect size against standard error and plotted it as a funnel plot to assess asymmetry visually. The significant asymmetry indicated the possibility of publication bias or heterogeneity. The meta-analysis was performed using Revman 5.3 (Cochrane Collaboration, Oxford, UK).

## Results

### Systematic Review

A systematic search identified 3,682 potential English articles, 1,957 were removed due to duplication. Two reviewers assessed the title and abstracts of 1,725 studies which 144 manuscripts remained for full-text assessment. Eventually, 18 RCTs were met the inclusion criteria. 2 RCTs were considered the same population of the TLIF group therefore one study was excluded from the analysis. The studies which did not report the variation were excluded. A preferred reporting Items for Systematic Reviews and Meta-Analyses diagram is shown in Fig. 1.

There were 15 RCTs included with 915 patients (470 TLIF, 258 PLF, 87 PLIF, 26 ALIF, 29 XLIF, 45 no fusion). The TLIF group in the 2 studies was in addition to posterolateral fusion (PLF). Publication years ranged from 2013 to 2019. Two studies reported outcome 1-year follow-up the other reported at least 2-year follow-up. Study characteristics are provided in Table 1.

**Table 1.** Study characteristics

| Author | Surgical technique | TLIF, n | Other techniques, n | Follow up |
| --- | --- | --- | --- | --- |
| Challier 2017 | PLF+TLIF vs PLF | 30 | 30 | 2y |
| Christensen<br>2014 | TLIF vs PLF | 51 | 49 | 1y, 2y |
| ElShazly 2013 | Discectomy+TLIF vs<br>no fusion | 15 | 15 | 2y |
|  | Discectomy+TLIF vs<br>Discectomy+PLF | 15 | 15 | 2y |
| Etemadifar<br>2016 | PLF+TLIF vs PLF | 25 | 25 | 1.5m, 3m, 6m,<br>1y, 2y |
| Fariborz 2016 | TLIF vs PLIF | 30 | 30 | 6m, 1y |
|  | TLIF vs PLF | 30 | 30 | 6m, 1y |
|  | TLIF vs no<br>fusion+instrumentation | 30 | 30 | 6m, 1y |
| Hoff 2016 | TLIF vs ALIF | 24 | 26 | 1y, 3y |
| Høy 2013* | TLIF vs PLF | 51 | 49 | 1y, 2y |
| Høy 2017* | TLIF vs PLF | 44 | 44 | 1y, 2y, 5-10y |
| Høy 2019* | TLIF vs PLF | 51 | 49 | 1y, 2y |
| Isaacs 2016** | TLIF vs XLIF | 26 | 29 | 1y, 2y |
| Jalalpour 2015 | TLIF vs PLF | 68 | 67 | 1y, 2y |
| Li 2013 | TLIF vs PLF | 19 | 18 | 2-5y |
| Putzier 2016 | TLIF vs PLIF | 24 | 23 | 1y |
| Sembrano<br>2016** | TLIF vs XLIF | 55 | 26 | 1y |
| Yang 2016 | TLIF vs PLIF | 32 | 34 | 3m, 1-2y |

\* same sample group, \*\* same sample group

### Quality assessment

For the risk of bias assessment, the included randomized controlled trials had a relatively high percentage of low risk in the randomization process and deviations from intended intervention domains. All included RCTs had some concerns risk of bias on measurement of the outcome. There was some high risk of bias in deviations from intended interventions and selection of the reported result domains. Detailed risk-of-bias assessment for included randomized controlled trials was provided in Fig. 2. A summary of the percentages of RCTs which were at low, some concerns, and high risk for each risk of bias domain wwaspresented in Fig. 3. The funnel plots showed no significant asymmetry which highlighted no evidence of publication bias on the fusion rate, total adverse events, and revision rate. (Supplementary File 1).

### Meta-analysis

A total of 15 included studies were included in the meta-analysis with 915 patients (470 TLIF, 258 PLF, 87 PLIF, 26 ALIF, 29 XLIF, 45 no fusion).

### Fusion rate

Fusion rate was 72.7% on TLIF group at 1 year follow up whereas 87.03% fusion rate was reported on other techniques. TLIF had slightly lower fusion rate at 1 year follow up compared to other techniques (RR=0.84 [95% CI 0.72, 0.97], *p*=0.02, *I*^*2*^=0.0%) (Fig. 4). However, the fusion rate at 2 years was not shown any statistically significant differences (RR=1.06 [95% CI 0.96, 1.18], *p*=0.27, *I*^*2*^=69.0%) as shown in Fig. 5.

**Fig. 4.**
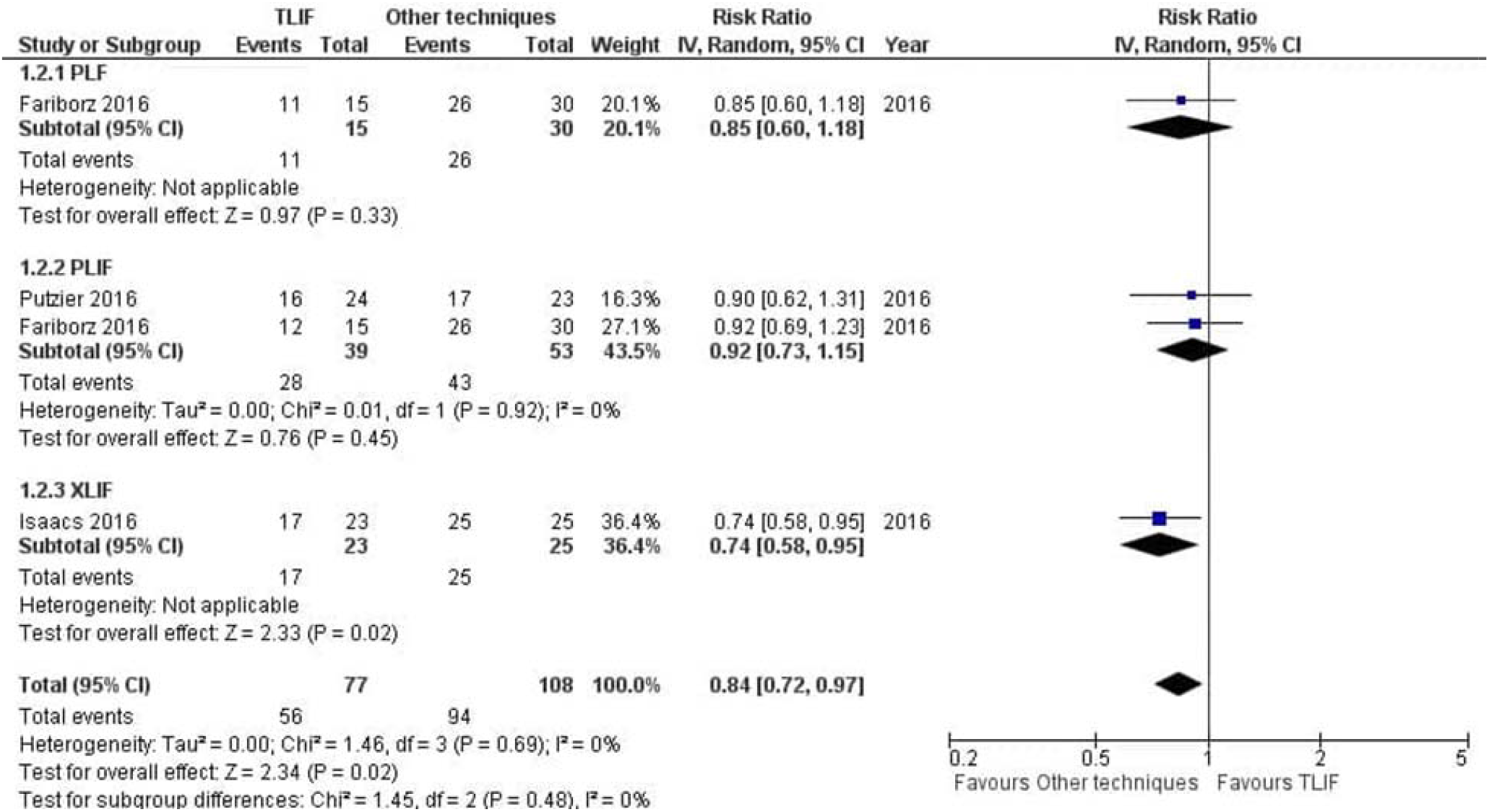
Forest plot and tabulated data illustrating the risk ratio (RR) for fusion rate at 1 year between TLIF, PLF, PLIF and XLIF showing that Other techniques had a better arm fusion rate at 1 year and was therefore superior to TLIF in this respect. *CI* confidence interval; *df* degrees of freedom

**Fig. 5.**
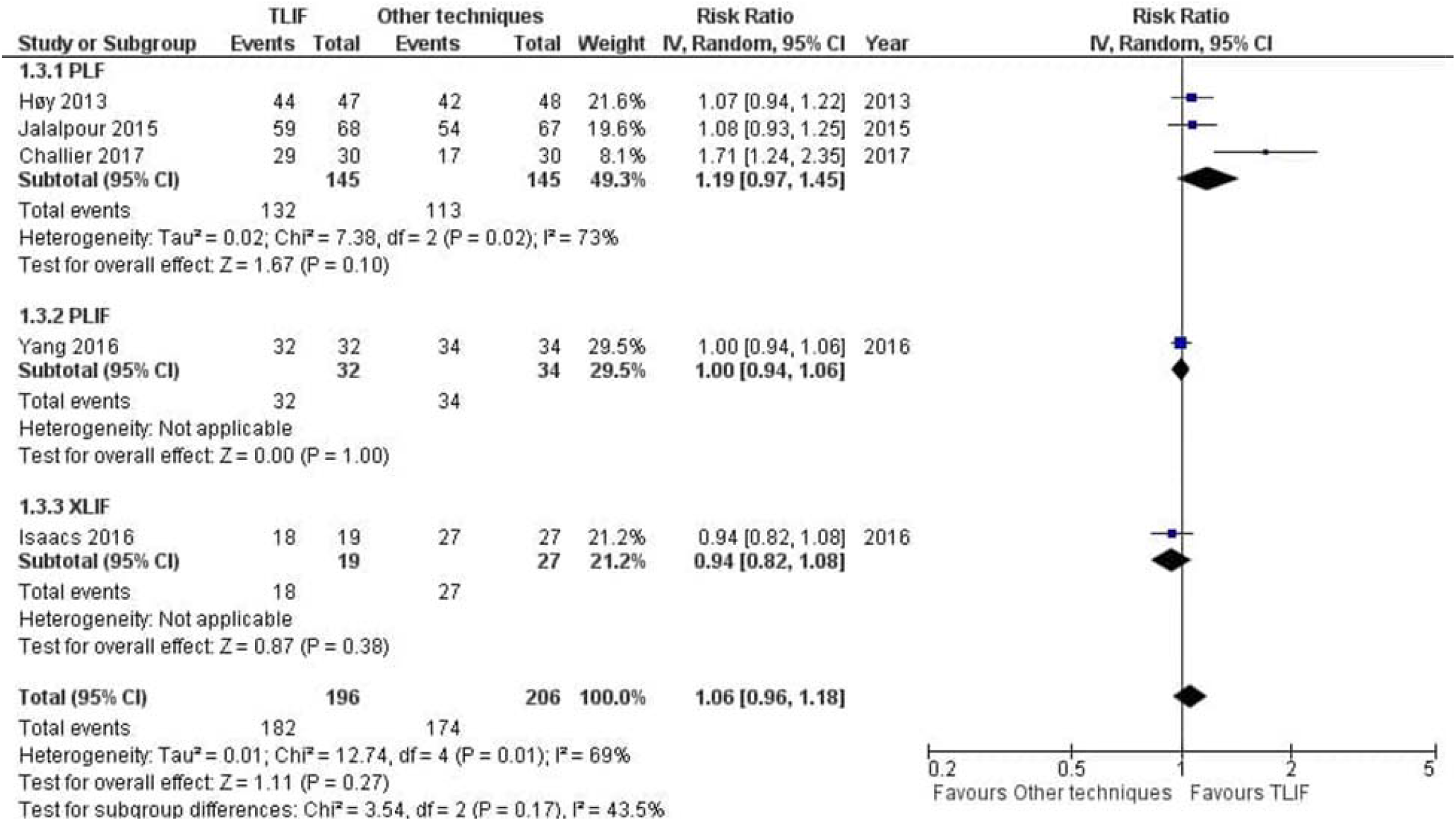
Forest plot and tabulated data illustrating the risk ratio (RR) for fusion rate at 2 years between TLIF, PLF, PLIF and XLIF showing that there was no significant difference of fusion rate at 2 years between procedures. *CI* confidence interval; *df* degrees of freedom

### Complications: total adverse events, revision, infection, and dural tear

Total adverse events were reported in 10 studies. TLIF had similar total adverse events compared with PLIF, XLIF and no fusion group (RR=0.90 [95% CI 0.59, 1.38], *p*=0.63, *I*^*2*^=0.0%) as shown in Fig. 6. For the revision needed after surgical procedures, the results indicated a different revision rate among groups. (RR=0.78 [95%CI 0.34, 1.79], *p*=0.56, *I*^*2*^=39.0%) as shown in Fig. 7.

**Fig. 6.**
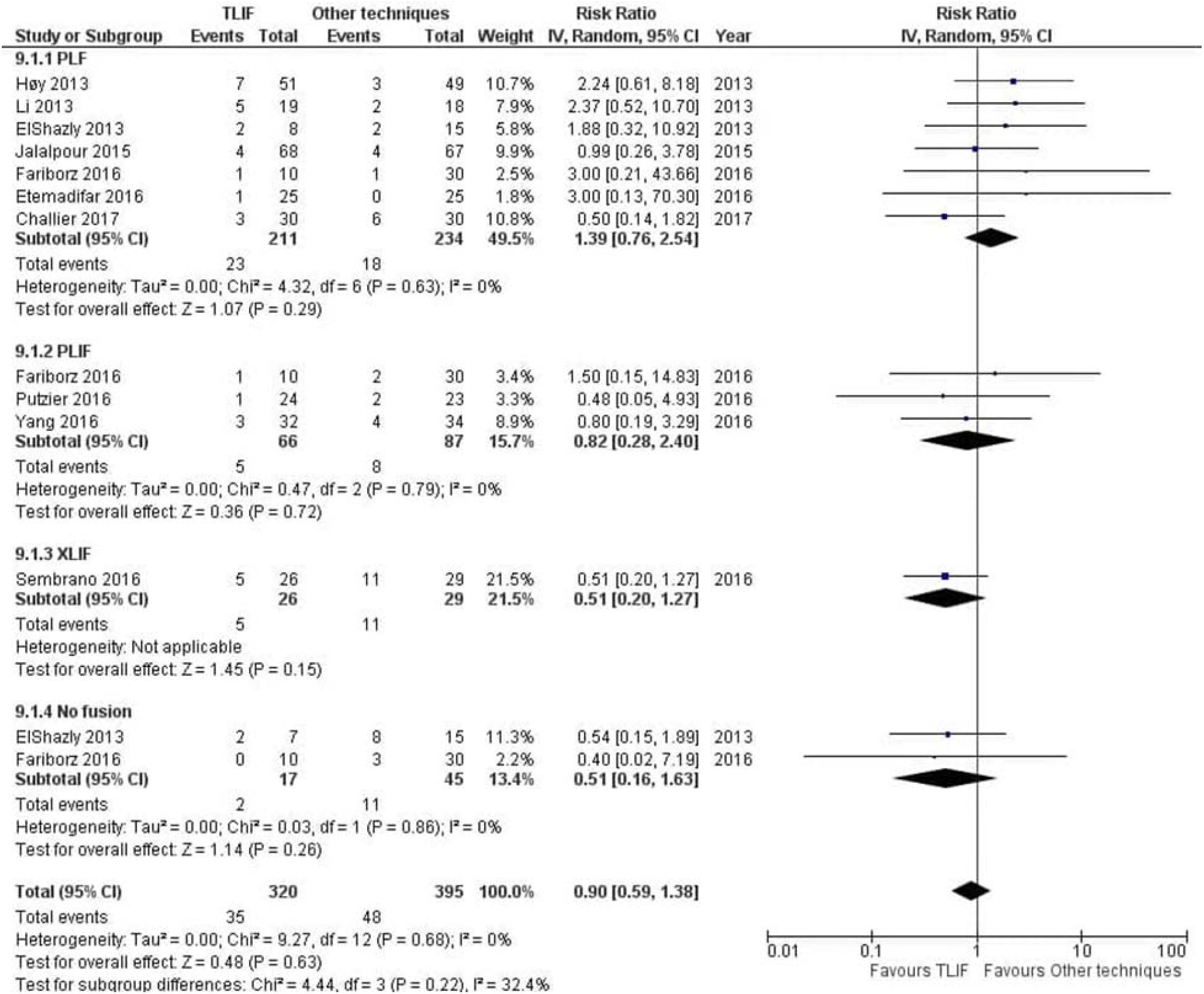
Forest plot and tabulated data illustrating the risk ratio (RR) for adverse events between TLIF, PLF, PLIF, XLIF and no fusion showing that there was no significant difference of adverse events between procedures. *CI* confidence interval; *df* degrees of freedom

**Fig. 7.**
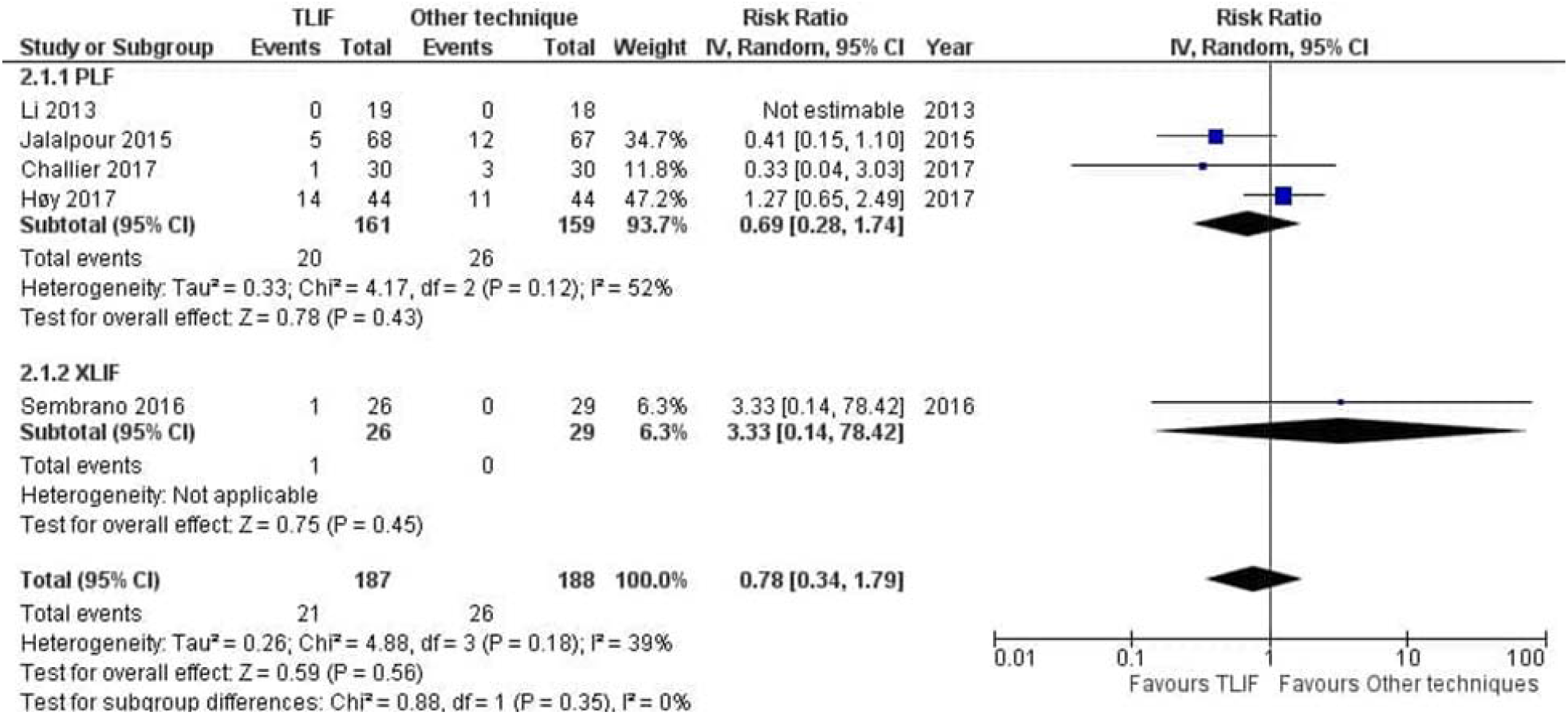
Forest plot and tabulated data illustrating the risk ratio (RR) for revision rate between TLIF, PLF and XLIF showing that there was no significant difference of revision rate between procedures. *CI* confidence interval; *df* degrees of freedom

Infection was reported in 6 studies, overall infection was similar among groups (RR=1.78 [95%CI 0.58, 5.46], *p*=0.31, *I*^*2*^=0.0%). More infection reported in the TLIF group but was not statistically significant. The dural tear was higher in other techniques esp. XLIF group but not reach statistically significant (RR=1.19 [95% CI 0.49, 2.89], *p*=0.70, *I*^*2*^=0.0%). The results of secondary outcomes were reported as shown in Table 2.

**Table 2.**
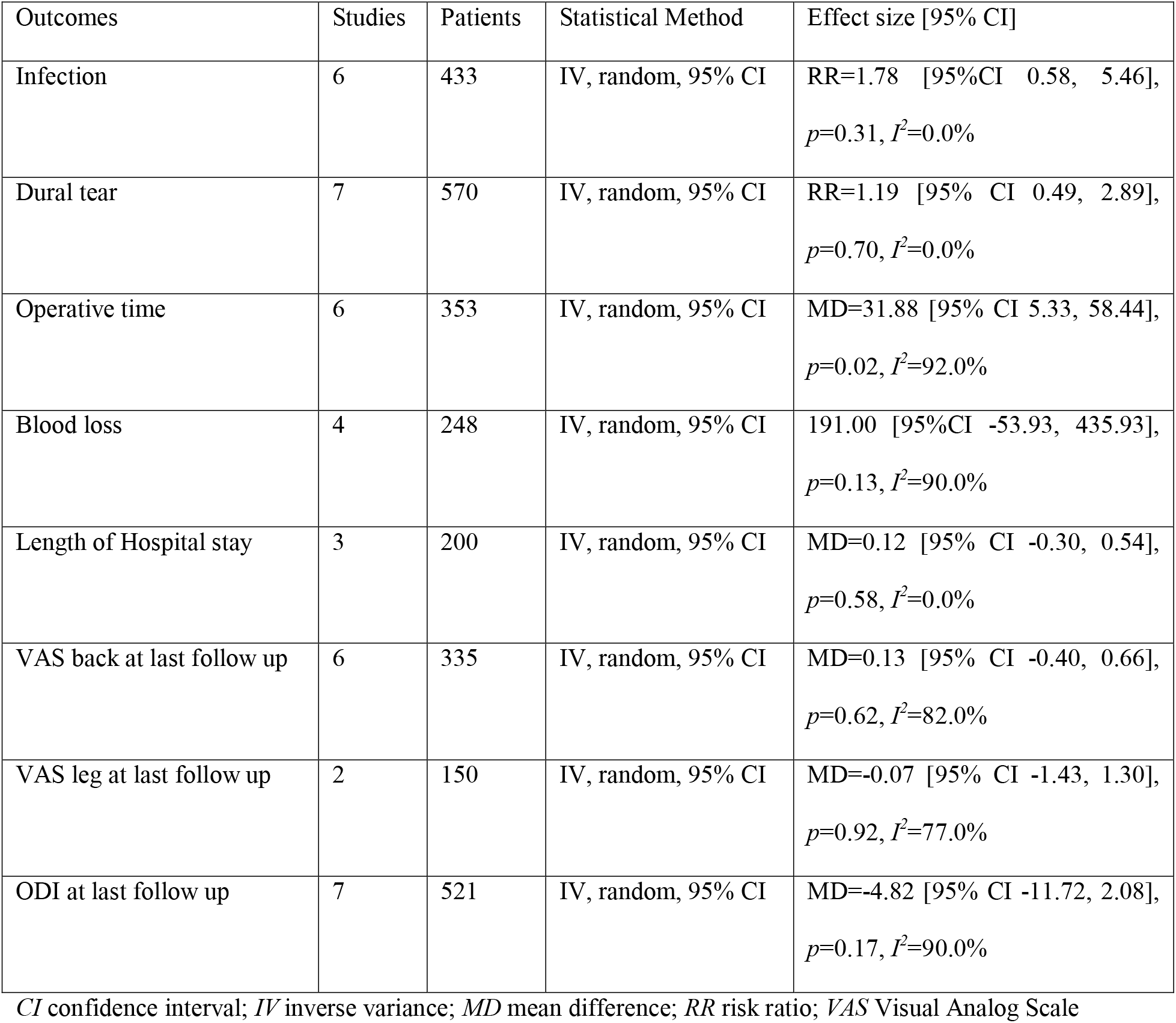
Secondary outcomes

| Outcomes | Studies | Patients | Statistical Method | Effect size [95% CI] |
| --- | --- | --- | --- | --- |
| Infection | 6 | 433 | IV, random, 95% CI | RR=1.78 [95% CI 0.58, 5.46],<br>$p=0.31$, $I^2=0.0\%$ |
| Dural tear | 7 | 570 | IV, random, 95% CI | RR=1.19 [95% CI 0.49, 2.89],<br>$p=0.70$, $I^2=0.0\%$ |
| Operative time | 6 | 353 | IV, random, 95% CI | MD=31.88 [95% CI 5.33, 58.44],<br>$p=0.02$, $I^2=92.0\%$ |
| Blood loss | 4 | 248 | IV, random, 95% CI | 191.00 [95% CI -53.93, 435.93],<br>$p=0.13$, $I^2=90.0\%$ |
| Length of Hospital stay | 3 | 200 | IV, random, 95% CI | MD=0.12 [95% CI -0.30, 0.54],<br>$p=0.58$, $I^2=0.0\%$ |
| VAS back at last follow up | 6 | 335 | IV, random, 95% CI | MD=0.13 [95% CI -0.40, 0.66],<br>$p=0.62$, $I^2=82.0\%$ |
| VAS leg at last follow up | 2 | 150 | IV, random, 95% CI | MD=-0.07 [95% CI -1.43, 1.30],<br>$p=0.92$, $I^2=77.0\%$ |
| ODI at last follow up | 7 | 521 | IV, random, 95% CI | MD=-4.82 [95% CI -11.72, 2.08],<br>$p=0.17$, $I^2=90.0\%$ |

*CI* confidence interval; *IV* inverse variance; *MD* mean difference; *RR* risk ratio; *VAS* Visual Analog Scale

### Operative time

ALIF, PLF, no fusion group has shorter operative time whereas PLIF has longer operative time compared to TLIF. The pooled mean difference in operative time of other techniques was 31.88 minutes shorter than TLIF (MD=31.88 [95% CI 5.33, 58.44], *p*=0.02, *I*^*2*^=92.0%)

### Blood loss

TLIF has less blood loss than PLIF 88.80 ml. No fusion has less blood loss among groups. Pooled mean difference in blood loss showed no significant difference (MD=191.00 [95%CI -53.93, 435.93], *p*=0.13, *I*^*2*^=90.0%).

### Length of hospital stay

Length of hospital stay between subgroup were not significantly difference. Pooled mean difference in hospital stay was 0.12. (MD=0.12 [95% CI -0.30, 0.54], *p*=0.58, *I*^*2*^=0.0%).

### Back and leg pain

Visual Analog Scale (VAS) for back were extracted from 6 studies. There was no difference between back pain at last follow up in TLIF and other techniques group (MD=0.13 [95% CI -0.40, 0.66], *p*=0.62, *I*^*2*^=82.0%). ALIF (MD=1.20 [95% CI 0.53, 1.87], *p*<0.01) and no fusion techniques (MD=0.60 [95% CI 0.08, 1.12], *p*=0.02) were shown less back pain at last follow up. Visual Analog Scale (VAS) for leg was extracted from only PLF studies. There was no difference between leg pain at last follow up in TLIF and PLF group (MD=-0.07 [95% CI -1.43, 1.30], *p*=0.92, *I*^*2*^=77.0%).

### ODI

No difference in ODI was observed (MD=-4.82 [95% CI -11.72, 2.08], *p*=0.17, *I*^*2*^=90.0%). Compared to TLIF, no fusion group had higher ODI at last follow up (MD=-41.30 [95% CI -50.15, -32.45], *p*<0.001).

## Discussion

Patients with degenerative lumbar spine disease require surgical intervention when the conservative treatments failed [7–10]. The operative methods are varyingly selected among spine surgeons. Therefore, the clinical outcomes esp. fusion rate, and other outcomes were reported in different studies. This systematic review and meta-analysis are to investigate the benefits and risks of lumbar interbody fusion, no fusion, and posterolateral fusion by comparing the fusion rate, clinical outcomes (VAS back, VAS leg, and ODI), parameters concerning operation (operative time, length of hospital stay, blood loss, revision) and complications in TLIF and other techniques for lumbar diseases.

Findings from our study were similar to the previous systematic review that reported 89.1% fusion rate and 12.5% reoperation rate [33]. Manzur et al. reported 85.6% fusion rate on LLIF [34]. Similar to Tao et al. study which PLIF compared with TLIF, demonstrated similar outcomes and complications [35].

Surgical complications evaluated by total adverse events were not shown statistically significant differences among lumbar interbody fusion, no fusion, and posterolateral fusion. However, TLIF seems to be safer than PLIF and ALIF in neural, spinal and vascular events, similar to previous study by Chi et al. [36]. Nonetheless, Yavin et al. demonstrated more complications on fusion group compared to non-fusion group [37].

### Strength and limitations

The strength of this study was that we included only RCTs that showed no significant asymmetry which highlighted no evidence of publication bias on the fusion rate, total adverse events, and revision rate. However, the small number of RCT on TLIF was the limitation of our study. The heterogeneity of the enrolled studies was another limitation. Furthermore, sample group of some the same population as shown in table 1. We try to reduce the bias by excluded the repeated data from the analysis.

The overall lumbar spine disease were heterogenous in approach for each studies which may complicated the results. Different surgical approach, experience and outcome measure were also observed. The outcomes were referred to single level surgery which couldn’t apply to multi-level diseases.

### Conclusion

The present systematic review and meta-analysis of RCTS demonstrated lower fusion rate of TLIF at 1 year but similar outcomes of TLIF with other techniques (no fusion, PLF, PLIF, ALIF, XLIF) for lumbar diseases in regards to fusion rate at 2 years, clinical outcomes and complications. Anticipate in randomized control trials are needed for selecting a surgical approach for lumbar spinal diseases.

## Supporting information

PRISMA 2009 Checklist

## Data Availability

All data generated or analyzed during this study are included in this published article (and its supplementary information files).

## Authors’ contribution

KW and KP contributed to the study conception and design. Material preparation and data collection were performed by KW and TN. Data analysis was performed by TN. All authors wrote the first draft of the manuscript, revised, and approved the final manuscript.

