## Supplementary Files for "Surgical Treatments for Lumbar Spine Diseases: A Systematic Review and Meta-Analysis"

**Supplementary Materials**

**1. Supplementary Table S1. Full search strategy**

**2. Supplementary Figure S1. Funnel plots: estimated prevalence and case fatality rate with standard error of each included study in meta-analysis**

**3. Supplementary Figure S2. Forest plots: secondary outcomes**

**Supplementary Table S1A** Full search strategy for PubMed

| Set # | PubMed | Results |
| --- | --- | --- |
| 1  TLIF | (transforaminal[tiab] AND (lif[tiab] OR (lumbar[tiab] AND interbody[tiab] AND (fusion[tiab] OR fusions[tiab])))) OR "transforaminal lif"[tiab] OR "transforaminal lumbar interbody fusion"[tiab] OR "transforaminal lumbar interbody fusions"[tiab] OR tlif[tiab] | **1540** |
| 2  RCT | "randomized controlled trial"[pt] OR "controlled clinical trial"[pt] OR trial[tiab] OR trials[tiab] OR random*[tiab] OR randomize[tiab] OR randomized[tiab] OR randomization[tiab] OR randomise[tiab] OR randomised[tiab] OR randomisation[tiab] OR randomly[tiab] OR placebo*[tiab] OR control*[tiab] OR controlled[tiab] OR ((single*[tiab] OR double*[tiab] OR treble*[tiab] OR triple*[tiab]) AND (blind*[tiab] OR mask*[tiab] OR masked[tiab])) OR allocat*[tiab] OR allocate[tiab] OR allocation[tiab] OR group[tiab] OR groups[tiab] OR "drug therapy"[sh] | **8449768** |
| 3 | #1 AND #2 | **714** |
| 4 | animals[MeSH Terms] NOT humans[MeSH Terms] | **4667834** |
| 5 | #3 NOT #4 | **707** |
| 6 | English[lang] | **25934721** |
| 7 | #5 AND #6 | **637** |

Last search 13 February 2020

**Supplementary Table S1B** Full search strategy for Embase

| Set # | Embase | Results |
| --- | --- | --- |
| 1  TLIF | ('transforaminal':ti,ab AND ('lif':ti,ab OR ('lumbar':ti,ab AND 'interbody':ti,ab AND ('fusion':ti,ab OR 'fusions':ti,ab)))) OR 'transforaminal lif':ti,ab OR 'transforaminal lumbar interbody fusion':ti,ab OR 'transforaminal lumbar interbody fusions':ti,ab OR 'tlif':ti,ab | **1980** |
| 2  RCT | 'randomized controlled trial'/exp OR 'double blind procedure'/exp OR 'single blind procedure'/exp OR 'crossover procedure'/exp OR 'controlled study'/de OR 'clinical trial'/de OR 'trial$':ti,ab OR 'random*':ti,ab OR 'placebo*':ti,ab OR 'control*':ti,ab OR (('singl$':ti,ab OR 'doubl$':ti,ab OR 'trebl$':ti,ab OR 'tripl$':ti,ab) AND ('blind*':ti,ab OR 'mask*':ti,ab)) OR 'allocat*':ti,ab OR 'group$':ti,ab OR 'crossover$':ti,ab OR 'cross over':ti,ab OR 'cross overs':ti,ab OR 'assign*':ti,ab OR 'compar*':ti,ab OR 'prospective*':ti,ab | **15901589** |
| 3 | #1 AND #2 | **1499** |
| 4 | [animals]/lim NOT [humans]/lim | **5747653** |
| 5 | #3 NOT #4 | **1480** |
| 6 | english:la | **30807902** |
| 7 | #5 AND #6 | **1372** |

Last search 13 February 2020

**Supplementary Table S1C** Full search strategy for Scopus

| Set # | Scopus | Results |
| --- | --- | --- |
| 1  TLIF | TITLE-ABS-KEY((transforaminal AND (lif OR (lumbar AND interbody AND (fusion OR fusions)))) OR "transforaminal lif" OR "transforaminal lumbar interbody fusion" OR "transforaminal lumbar interbody fusions" OR tlif) | **1687** |
| 2  RCT | TITLE-ABS-KEY(trial OR trials OR random* OR placebo* OR control* OR ((single* OR double* OR treble* OR triple*) AND (blind* OR mask*)) OR allocat* OR group*) | **21299454** |
| 3 | #1 AND #2 | **901** |
| 4 | ALL(animals AND NOT humans) | **3927099** |
| 5 | #3 AND NOT #4 | **896** |
| 6 | LANGUAGE(english) | **66776799** |
| 7 | #5 AND #6 | **796** |

Last search 13 February 2020

**Supplementary Table S1D** Full search strategy for Web of Science

| Set # | Web of Science | Results |
| --- | --- | --- |
| 1  TLIF | TS=((transforaminal AND (lif OR (lumbar AND interbody AND (fusion OR fusions)))) OR "transforaminal lif" OR "transforaminal lumbar interbody fusion" OR "transforaminal lumbar interbody fusions" OR tlif) | **1328** |
| 2  RCT | TS=(trial OR trials OR random* OR placebo* OR control* OR ((single* OR double* OR treble* OR triple*) AND (blind* OR mask*)) OR allocat* OR group*) | **9432516** |
| 3 | #1 AND #2 | **632** |
| 4 | ALL=(animal NOT human) | **1059498** |
| 5 | #3 NOT #4 | **627** |
| 6 | #5 restrict language to English | **617** |

Last search 13 February 2020

**Supplementary Table S1E** Full search strategy for CENTRAL

| Set # | CENTRAL | Results |
| --- | --- | --- |
| 1  TLIF | (transforaminal:ti,ab,kw AND (lif:ti,ab,kw OR (lumbar:ti,ab,kw AND interbody:ti,ab,kw AND (fusion:ti,ab,kw OR fusions:ti,ab,kw)))) OR "transforaminal lif":ti,ab,kw OR "transforaminal lumbar interbody fusion":ti,ab,kw OR "transforaminal lumbar interbody fusions":ti,ab,kw OR tlif:ti,ab,kw | **260** |
| 2 | [mh animals] NOT [mh humans] | **7304** |
| 3 | #1 NOT #2 | **260** |

Last search 13 February 2020

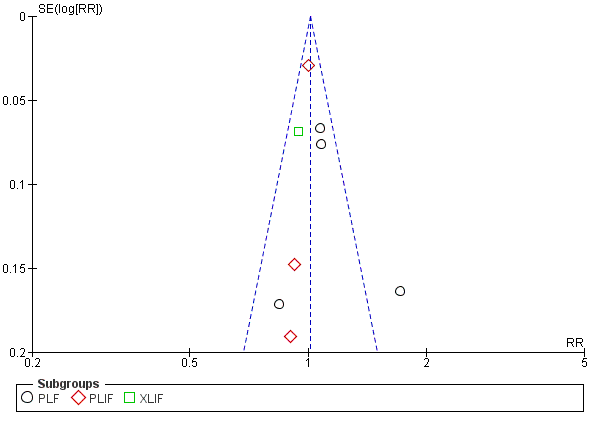

**Supplementary Fig. S1A** Funnel plot: estimated relative risk of fusion rate at last follow up and standard error of each included study in meta-analysis. Black circles represent estimated relative risk for included studies with PLF as comparator. Red diamonds represent estimated relative risk for included studies with PLIF as comparator. Green square represents estimated relative risk for included study with XLIF as comparator. Vertical blue line represents fixed effect estimated relative risk. Sides of the triangle represent the expected 95% confidence intervals with inverted funnel shape.

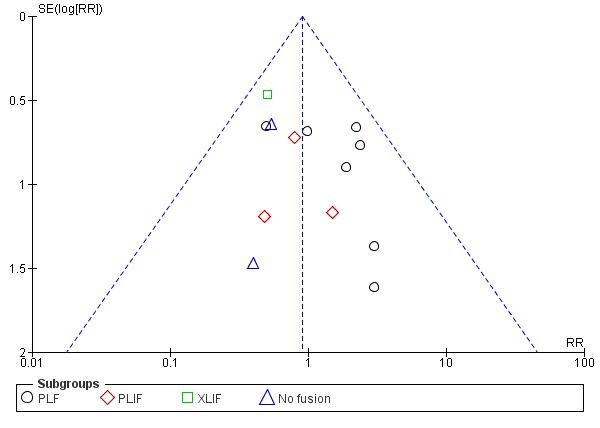

**Supplementary Fig. S1B** Funnel plot: estimated relative risk of total adverse events and standard error of each included study in meta-analysis. Black circles represent estimated relative risk for included studies with PLF as comparator. Red diamonds represent estimated relative risk for included studies with PLIF as comparator. Green square represents estimated relative risk for included study with XLIF as comparator. Blue triangles represent estimated relative risk for included study with No fusion as comparator. Vertical blue line represents fixed effect estimated relative risk. Sides of the triangle represent the expected 95% confidence intervals with inverted funnel shape.

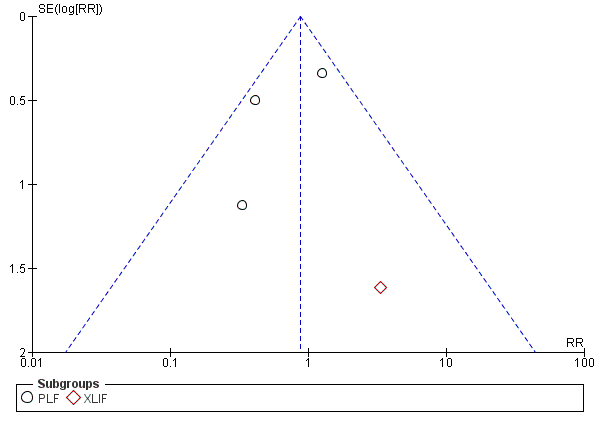

**Supplementary Fig. S1C** Funnel plot: estimated relative risk of fusion rate at last follow up and standard error of each included study in meta-analysis. Black circles represent estimated relative risk for included studies with PLF as comparator. Red diamonds represent estimated relative risk for included studies with PLIF as comparator. Vertical blue line represents fixed effect estimated relative risk. Sides of the triangle represent the expected 95% confidence intervals with inverted funnel shape.

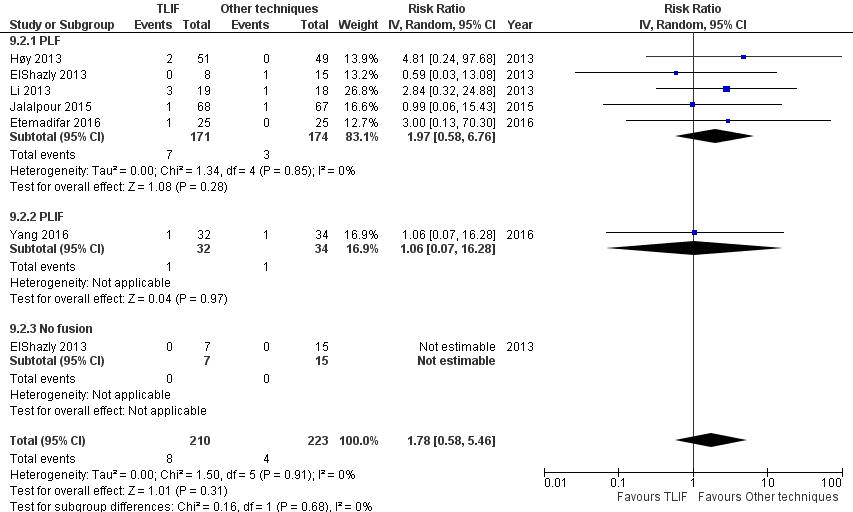

**Supplementary Fig S2A** Forest plot and tabulated data illustrating the risk ratio (RR) for infection rate between TLIF, PLF, PLIF and no fusion showing that there was no significant difference of infection rate between procedures. *CI* confidence interval; *df* degrees of freedom

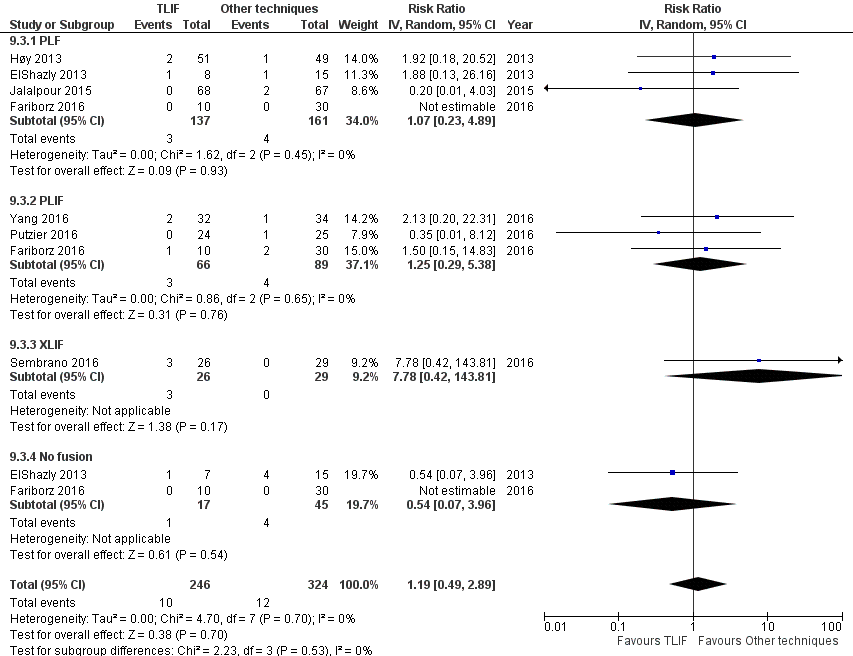

**Supplementary Fig S2B** Forest plot and tabulated data illustrating the risk ratio (RR) for dural tear between TLIF, PLF, PLIF, XLIF and no fusion showing that there was no significant difference of dural tear between procedures. *CI* confidence interval; *df* degrees of freedom

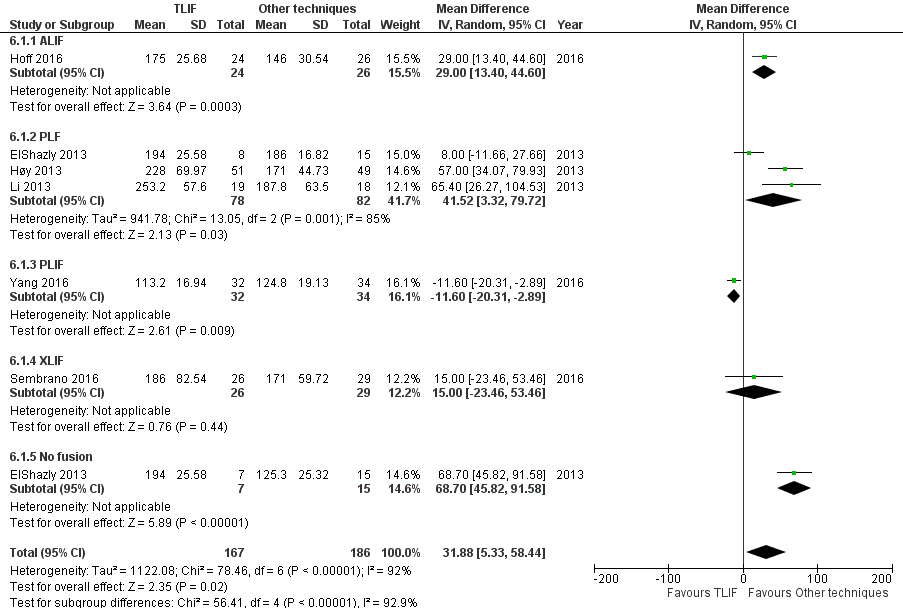

**Supplementary Fig S2C** Forest plot and tabulated data illustrating the mean difference for operative time between TLIF, PLF, PLIF, XLIF and no fusion showing that operative time of other techniques was significantly shorter than TLIF. *CI* confidence interval; *df* degrees of freedom

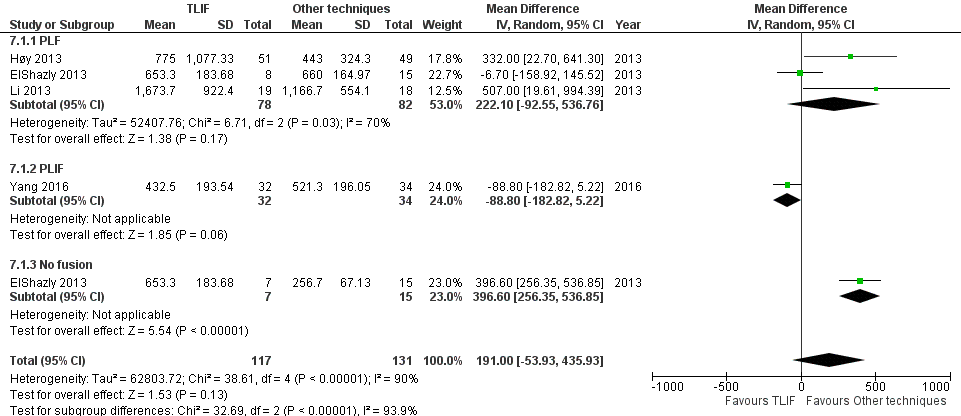

**Supplementary Fig S2D** Forest plot and tabulated data illustrating the mean difference for blood loss between TLIF, PLF, PLIF and no fusion showing that there was no significant difference of blood loss between procedures. *CI* confidence interval; *df* degrees of freedom

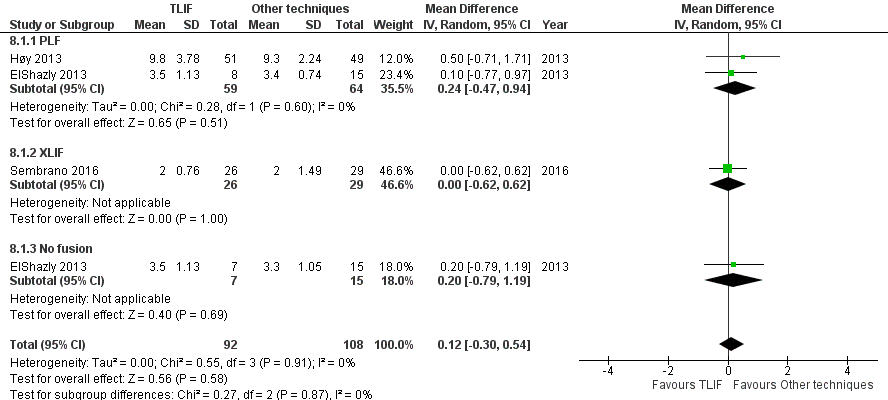

**Supplementary Fig S2E** Forest plot and tabulated data illustrating the mean difference for length of hospital stay between TLIF, PLF, XLIF and no fusion showing that there was no significant difference of length of hospital stay between procedures. *CI* confidence interval; *df* degrees of freedom

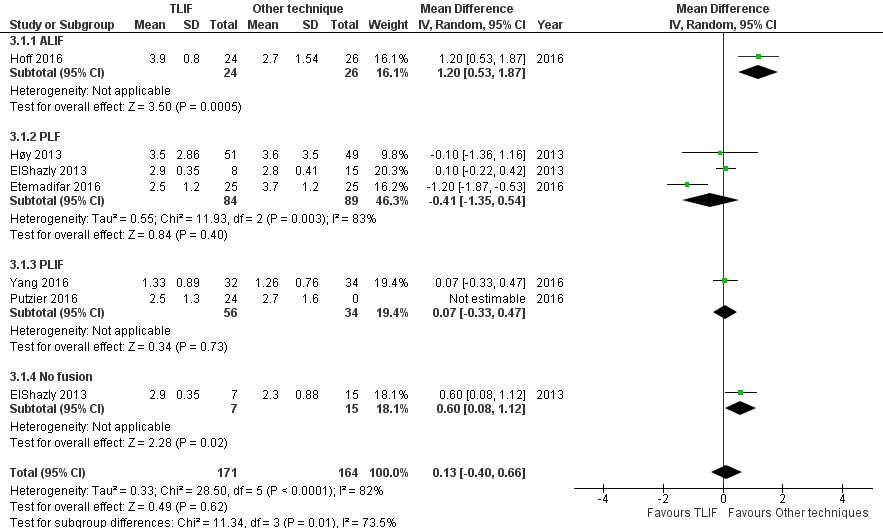

**Supplementary Fig S2F** Forest plot and tabulated data illustrating the mean difference for back pain at last follow up between TLIF, ALIF, PLF, PLIF and no fusion showing that there was no significant difference of back pain at last follow up between procedures. *CI* confidence interval; *df* degrees of freedom

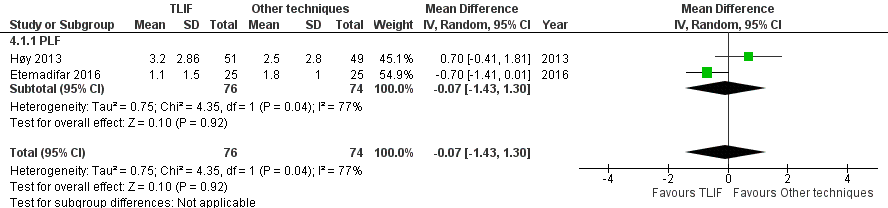

**Supplementary Fig S2G** Forest plot and tabulated data illustrating the mean difference for leg pain at last follow up between TLIF and PLF showing that there was no significant difference of leg pain at last follow up between procedures. *CI* confidence interval; *df* degrees of freedom

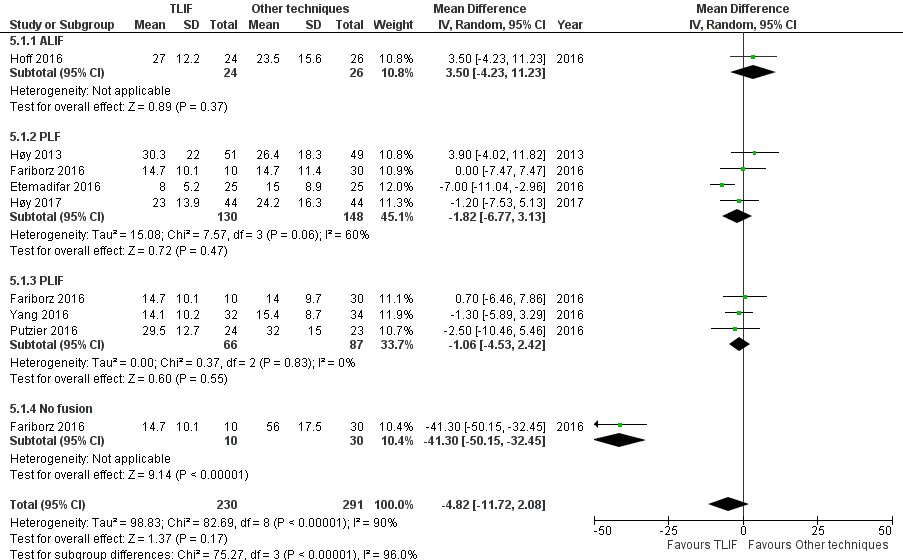

**Supplementary Fig S2G** Forest plot and tabulated data illustrating the mean difference for ODI at last follow up between TLIF, ALIF, PLF, PLIF and no fusion showing that there was no significant difference of ODI at last follow up between procedures. *CI* confidence interval; *df* degrees of freedom
